# Psychometric properties of the Thai version of the Illness-Specific Social Support Scale Short Version-8 (ISSS-8) among hematological malignancy patients in the Northeastern region of Thailand: A multicenter study

**DOI:** 10.64898/2026.01.15.26344141

**Authors:** Ueamporn Summart, Monthida Sangruangake, Chaliya Wamaloon, Acharaporn Moungmultri, Muhamad Zulfatul A’la

## Abstract

Social support, which is an essential aspect influencing health, has resulted in the development of several approaches for assessment in cancer patients. The Illness-Specific Social Support Scale Short Version-8 (ISSS-8) effectively evaluates social support among diverse patient demographics; however, its psychometric validity has yet to be established in Asian cultural contexts. The objectives of this study were to translate and culturally adapt the ISSS-8 for the Thai setting and to assess its psychometric properties in patients with hematological malignancies. This study employed a convenience sampling method to select patients with hematological malignancies undergoing hospitalization at three tertiary institutions in Northeastern Thailand. Psychometric testing was conducted following the translation and cross-cultural adaptation of the ISSS-8 into Thai. A total of 350 patients were recruited. Participants were randomly divided into two groups for exploratory factor analysis (EFA) (n = 200) and confirmatory factor analysis (CFA) (n = 150). The exploratory factor analysis of the eight items yielded a loading from a two-factor model comprising Positive Support and Detrimental Interactions, which explained 79.09% of the variance. Cronbach’s alpha (0.80) and item–total correlations (rho = 0.33–0.65) demonstrated acceptable reliability of the ISSS-8, while test–retest reliability was high (ICC = 0.924–0.934). The average variance extracted (AVE) demonstrated convergent validity for all ISSS-8 subscales, with AVEs ranging from 0.76 to 0.80. Moreover, this instrument demonstrated a significant positive correlation with the Stanford Inventory of Cancer Patient Adjustment (r = 0.244). The ISSS-8 is a short, accurate, and valid instrument for measuring social support in Thai patients with hematological malignancies. As a result, healthcare professionals can use the ISSS-8 to assess social support in both research and clinical settings. The findings underscore the importance of integrating social support evaluation into cancer management and family-centered care, thereby supporting comprehensive and adaptive healthcare delivery in Thailand.

## Introduction

Hematological malignancies (HMs), including leukemia and lymphoma, pose a considerable global health challenge, with incidence rates rising since 1990, culminating in approximately 1.34 million cases in 2019 [1]. Chemotherapy is the primary and indispensable treatment for the majority of hematological malignancies due to the systemic nature of the disease and the accelerated growth rate inherent to malignant hematological cells [2]. Treatment for these types of malignancies usually involves long, intense chemotherapy regimens, which have been associated with extensive physical and mental distress, including extreme fatigue, anxiety, and depression, as well as a substantial decline in daily functioning and quality of life (QOL) [3]. Considering the severity and duration of treatment, it is crucial to employ appropriate psychological assessments to facilitate patient adjustment to illness and treatment and optimize clinical outcomes. Some cancer patients are able to cope with their diagnosis and treatment, whereas others experience negative emotional responses, including lethargy, fragility, melancholy, and anxiety, particularly at the initial stage [4]. These emotions are frequently followed by incapacitation, frailty, sadness, trauma, panic, and concerns over personal survival [5].

Social support is widely acknowledged as a crucial factor influencing psychological well-being, stress adaptation, and health outcomes among various populations [6]. It serves as a significant mediator of the relationship between disease and psychological adjustment for both patients and their family members [7]. Growing evidence indicates that adequate social support is associated with less emotional distress, improved coping strategies, and higher QOL, whereas inadequate or conflict-laden social support exacerbates stress and impedes adaptability [8]. In patients with hematological malignancies, who frequently undergo intensive therapies including chemotherapy, stem cell transplantation, and prolonged hospitalization, approximately 40–60% of patients experience increased mental distress [9]. Previous research indicates that cancer patients with substantial social support exhibit improved QOL and reduced mortality, whereas those lacking sufficient support experience inferior oncological outcomes, expedited disease progression, and diminished overall survival [10, 11]. Consequently, numerous researchers regard social support as a significant indicator of health, which has resulted in the development of various methods to assess social support in cancer patients.

In the Thai setting, the revised Thai Multidimensional Scale of Perceived Social Support (r-Thai MSPSS) is a commonly utilized instrument for assessing social support among cancer patients. It was developed and validated among medical students [12]. This tool offers advantages including brevity, established reliability, and comparability across studies [12, 13]. However, this instrument’s focus on broad, positively perceived help may limit its sensitivity to the complex, context-specific dimensions of social support experienced by cancer patients [14].

To address the limitations of the r-Thai MSPSS, the Illness-Specific Social Support Scale Short Version-8 (ISSS-8), a commonly employed abbreviated version of the original ISSS, is acknowledged as having strong internal consistency across its two subscales [15]. This tool was developed as an alternative instrument to measure social support and has been translated and validated for application in diverse populations, primarily focusing on patients with cancer or other chronic illnesses, predominantly in Western contexts [15–17]. Validation studies have confirmed a two-factor structure, comprising the Positive Support and Detrimental Interactions subscales, which demonstrate robust psychometric properties across various cancer groups, including melanoma patients [17].

The ISSS-8 accurately assesses social support across various patient demographics [17–19]; however, its psychometric validity remains unproven in both the general population and clinical settings within Asian cultural contexts. To date, no research has examined the ISSS-8 in Thailand. Validation studies of this scale are also limited, particularly among individuals with hematological malignancies. This research deficiency clearly fails to meet the requirements for practical applications. Thailand’s collectivist culture emphasizes family participation in disease care, potentially leading to different patterns of social support provision and reception compared to Western countries, where the ISSS-8 framework was initially developed [20]. This cultural difference may affect how social support is measured in Thailand, and, consequently, the accuracy and usefulness of the scale for this patient group [21].

Therefore, the ISSS-8’s factor structure, reliability, and validity require further assessment within this cohort to confirm that the Thai version accurately and consistently measures the underlying dimensions. Misinterpretation of support dynamics stemming from an unvalidated measure may result in inefficient or detrimental psychological therapies [22]. This study will enable healthcare practitioners in Thailand to more efficiently address the specific requirements of families by facilitating personalized care, while enhancing the overall quality of treatment. Practitioners can create a more supportive atmosphere for patients and their families by learning about the different cultural settings and support systems [23]. The rising incidence and prevalence of patients with hematological malignancies necessitate the validation of the psychometric properties of the ISSS-8 to ensure adequate social support through strategies designed to address patients’ psychosocial and interactive needs. The present study sought to translate and culturally adapt the ISSS-8 for the Thai context and evaluate its psychometric properties in patients with hematological malignancies, emphasizing content validity, construct validity, convergent validity, and reliability.

## Materials and methods

### Study design and participants

This study employed a multicenter, methodological design using data from a larger project, namely “Self-care behaviors and the relationship between health literacy, self-efficacy, social support, and self-care behaviors among patients with lymphoma receiving chemotherapy.” This study utilized a convenience sampling approach to select patients with hematological malignancies receiving treatment or hospitalized at three tertiary hospitals in the Northeastern region of Thailand from August 1 – December 31, 2025, using self-assessment questionnaires. The criteria for inclusion were: 1) a diagnosis of leukemia, lymphoma, or multiple myeloma and undergoing chemotherapy; 2) aged ≥ 18 years; 3) adequate reading and writing skills in Thai to complete the survey; and 4) willingness to participate. The criteria for exclusion were: 1) simultaneous treatment for other malignancies; 2) being in the terminal stage of the illness; and 3) insufficient cognitive or physical ability to participate in the study.

This study involved the translation and cultural adaptation of the ISSS-8 into Thai to assess social support among patients with hematological malignancies in Thailand. The original author authorized the translation and adaptation of the ISSS-8 for use in Thailand via email. The research received approval from the university’s ethics committee and was conducted in compliance with recognized ethical standards.

### Translation and cultural adaptation

The English version of ISSS-8 items was translated into Thai using Brislin’s forward and backward translation method as part of the questionnaire development [24]. Four English-Thai multilingual translators were identified: two translated the ISSS-8 into Thai, and the other two translated it back into English. Subsequently, two native English speakers compared the original ISSS-8 with the back-translated version. The Thai version of the ISSS-8 was then tested in a pilot group of 30 Thai patients with hematological malignancies to assess the translation’s effectiveness and the test’s usability. To ensure comprehension, participants were asked to read and listen to all items.

### Study instruments

The first section of the questionnaire was composed of patient demographic data, including gender, age, educational level, marital status, occupation, income, type of hematological malignancy, stage of cancer, and chemotherapy regimen.

The German adaptation of the Illness-Specific Social Support Scale Short Version-8 (ISSS-8) was executed by Ramm and Hasenbring [16] based on the original version by Revenson et al. [25]. The eight-item German version of the ISSS assesses patients’ self-reported positive and negative social support related to chronic illnesses. The detailed instructions for the ISSS-8 are: “These questions relate to your connections with key individuals: your partner, family members, friends and acquaintances, colleagues, and neighbors. We aim to understand your views and evaluations on these relationships. The behavior of others can be beneficial, occasionally detrimental, and at times, unpleasant. The following are behaviors exhibited by individuals when someone is unwell. Kindly indicate the frequency with which one or more of your acquaintances have exhibited such behavior towards you.” The ISSS-8 [15] has two subscales: ’Positive Support’ (four items) and ’Detrimental Interactions’ (four items), which assess positive and negative social support, respectively. Items are evaluated using a five-point Likert scale, with values from 0 (’never’) to 4 (’always’), resulting in a subscale score range of 0 to 16. On the ’Positive Support’ subscale, elevated values signify enhanced support, whereas higher scores on the ’Detrimental Interactions’ subscale imply more significant negative interactions between the patient and their loved ones [15]. Revenson et al. [25] assert that positive and negative interactions are independent of one another. The two subscales demonstrated internal consistencies with Cronbach’s alpha values of 0.88 and 0.68, respectively [16].

The Thai version of the Stanford Inventory of Cancer Patient Adjustment (SICPA) is a 38-item tool designed to assess women’s trust in their capacity to handle cancer-related challenges [26]. The SICPA total score ranges from 0 to 380, with elevated numbers signifying a greater level of self-efficacy. The baseline internal consistency of the SICPA in this study was satisfactory (Cronbach’s alpha = 0.87).

### Sample size calculation

The sample size for confirmatory factor analysis (CFA) and structural equation modeling (SEM) was determined using the N: q rule, representing the ratio of participants (N) to model parameters (q), in accordance with Kline’s recommendation of a 10:1 ratio [27]. Considering the number of items in the ISSS-8 (n = 8), a minimum required sample size of 80 participants was established. Consequently, 350 patients with hematological malignancies were included in this cross-sectional study. To mitigate model overfitting, exploratory factor analysis (EFA) and confirmatory factor analysis (CFA) were conducted on participants randomly allocated into two groups (group 1, n = 200; group 2, n = 150).

### Statistical analysis

All statistical analyses in this study were performed utilizing IBM SPSS and AMOS version 26. The demographic data of the participants were characterized using descriptive statistics, encompassing means and standard deviations for continuous variables, as well as frequencies and percentages for categorical data. Data were checked for missing values before analysis, and only fully answered questionnaires were included in the analysis. In addition, the skewness and kurtosis of all ISSS-8 items were initially evaluated to determine normality.

The evaluation of floor and ceiling effects was conducted by analyzing the proportion of respondents who achieved scores at the floor (minimum score) and ceiling (maximum score), respectively. We established that a floor or ceiling effect would be considered relevant based on the empirical threshold of 15%, and a cumulative ceiling or flooring of 50%, as proposed. Floor and ceiling effects can indicate that extreme items are missing in either end of the scale, which can possibly limit its validity [28]. Additionally, the discriminative capacity of the items was evaluated by corrected item–rest polyserial correlation, with acceptable indices exceeding 0.20 [29].

Parallel analysis (based on principal component analysis) was employed with sample group 1 (n = 130) to ascertain the number of components in the exploratory factor analysis (EFA) of the ISSS-8 measurement model. The factor structure was subsequently examined through principal axis factoring with varimax rotation. Factor loadings below 0.50 were considered negligible, and item cross-loadings exceeding 0.20 were sequentially removed [30]. Additionally, factor loadings were employed to compute the average variance extracted (AVE) and composite reliability (CR). After establishing the principal axis factoring results, construct validity was evaluated using CFA. The first-order two factorial model of the ISSS-8 and the second-order unique-factor model of the ISSS-8 were evaluated. A robust maximum likelihood estimator was utilized to address multivariate non-normality. Hu and Bentler’s criteria [31] for various fit indices were employed to assess the fit of the proposed model to the data. The chi-square test statistic was employed; however, due to its sensitivity to sample size, additional fit indices were assessed: (a) the comparative fit index (CFI ≥ 0.90 signifies a good fit); (b) the root mean square error of approximation (RMSEA ≤ 0.08 denotes an acceptable fit); and (c) the standardized root mean square residual (SRMR ≤ 0.08 indicates an adequate fit). A chi-square difference test, utilizing the Satorra–Bentler scaled chi-square test [32], was employed to compare the first-order two factorial model of the ISSS-8 with the second-order unique-factor model of social support. Along with the CFA, the Kaiser–Meyer–Olkin (KMO) measure of sampling adequacy and Bartlett’s test of sphericity were conducted to demonstrate factorability [30].

Reliability was evaluated through both CR and Cronbach’s alpha. CR is deemed suitable when the values for both subscales surpass 0.60 [33]. Internal consistency reliability was evaluated through Cronbach’s alpha, with values exceeding 0.70 for all subscales considered satisfactory. Item–total correlations were also examined, with a threshold of ≥ 0.30 to ensure that each item contributed meaningfully to its scale and to avoid redundancy [34].

The convergent validity of the ISSS-8 was evaluated based on AVE, which indicated the proportion of variance explained by each latent variable derived from all items within that variable. An AVE value exceeding 0.50 signifies that the latent variable exhibits adequate convergent validity [31]. Likewise, the square root of AVE for each sub-dimension should be greater than the correlation coefficients between sub-dimensions to verify discriminant validity [31].

Concurrent validity was measured using Pearson’s correlation coefficient. We hypothesized a positive correlation between the ISSS-8 and self-efficacy as measured by the SICPA, assuming that high correlation implies good concurrent validity and suggests that the two scales assess similar concepts [35]. Following Cohen’s criteria, the correlation between the variables was classified as small, medium, or large, with correlation coefficients greater than 0.10, 0.30, and 0.50, respectively [36].

Discriminant validity was assessed by comparing the square root of AVE with the correlation coefficient of observed variables. A square root of AVE greater than the correlation coefficient signifies stronger discriminant validity [37]. Test–retest reliability was assessed in a sub-sample of 30 participants to determine the intraclass correlation (ICC). Reliability was considered adequate when ICC values surpassed the threshold of 0.70 [34].

### Ethical considerations

The study followed the ethical standards and principles outlined in the Declaration of Helsinki. Ethical clearance for the study protocol was granted by the Roi Et Rajabhat University Ethics Committee for Human Research (Reference No. RERU-EC 058/2568), Roi Et Hospital (Reference No. RE 130/2568), Udon Thani Hospital (Reference No. UDH-REC 50/2568), and Ubon Ratchathani Cancer Hospital (Reference No. EC 025/2025). Each study participant provided informed written consent to participate in this investigation. Furthermore, each participant was assured of the confidentiality of his or her personal information.

### Data availability statement

All relevant data are contained within the manuscript and its supporting information files.

## Results

### General information of the participants

Participant characteristics are detailed in Table 1. Approximately 63.6% of the participants were female, with ages ranging from 19 to 91 years (M = 53.32; SD = 12.30). Over 47.7% of the participants had attained secondary education. Lymphoma was the predominant diagnosis, accounting for 72.6% of cases, with stage III comprising 35.4%.

**Table 1.** Demographic characteristics of patients with hematological malignancies (n = 350).

| Baseline characteristic | N | % |
| --- | --- | --- |
| Gender |  |  |
| Male | 256 | 73.1 |
| Female | 94 | 26.9 |
| Age (years) |  |  |
| Mean (SD) | 53.32 (12.30) |  |
| Median (min: max) | 54.0 (19:91) |  |
| Education level |  |  |
| Primary | 114 | 32.6 |
| Secondary | 167 | 47.7 |
| Bachelor's or higher | 69 | 19.7 |
| Religion |  |  |
| Buddhism | 296 | 84.6 |
| Christianity | 27 | 7.7 |
| Islam | 27 | 7.7 |
| Occupation |  |  |
| Agriculture | 141 | 40.2 |
| Civil servant | 14 | 5.2 |
| Employee | 118 | 32.6 |
| Personal business | 35 | 10.0 |
| Others | 42 | 12.0 |
| Household income (baht) |  |  |
| Less than 10,000 | 198 | 56.5 |
| 10,000-14,999 | 92 | 26.3 |
| 15,000-19,999 | 23 | 6.8 |
| ≥ 20,000 | 37 | 10.4 |
| Marital status |  |  |
| Single | 117 | 33.4 |
| Married | 230 | 65.7 |
| Divorced/separated | 3 | 0.9 |
| Cancer diagnosis |  |  |
| Lymphoma | 254 | 72.6 |
| Leukemia | 96 | 27.4 |
| Stage of cancer |  |  |
| I | 70 | 20.0 |
| II | 116 | 33.1 |
| III | 124 | 35.4 |
| IV | 30 | 8.6 |
| Unknown | 10 | 2.9 |

### Descriptive analysis and item analysis of the ISSS-8

The descriptive analysis of the ISSS-8 items is presented in Table 2. The average scores of the responses to the eight items from all participants ranged from 2.61 to 3.17. For the item analysis, the results showed that all items demonstrated acceptable normality, with skewness and kurtosis values less than ±1. Floor effects were observed for all Detrimental Interactions items, whereas ceiling effects were observed for all Positive Support items. In terms of discriminative capacity, all items had polyserial item–rest correlation coefficients greater than 0.20 **(Table 2)**.

**Table 2.** Distribution of responses and discrimination of the items.

| Item | Response (%) |  |  |  |  | Mean | SD | Item–rest correlation |
| --- | --- | --- | --- | --- | --- | --- | --- | --- |
|  | 0 | 1 | 2 | 3 | 4 |  |  |  |
| 1 | 0.0 | 4.9 | 11.2 | 49.2 | 34.6 | 3.17 | 0.06 | 0.40 |
| 2 | 4.4 | 0.5 | 20.0 | 46.3 | 28.8 | 3.07 | 0.07 | 0.48 |
| 3 | 0.0 | 4.9 | 11.2 | 49.3 | 34.6 | 3.17 | 0.06 | 0.40 |
| 4 | 3.9 | 3.9 | 16.6 | 46.8 | 26.3 | 3.13 | 0.06 | 0.57 |
| 5 | 31.7 | 12.2 | 38.5 | 14.6 | 2.9 | 3.39 | 0.08 | 0.52 |
| 6 | 32.7 | 11.7 | 32.7 | 15.1 | 7.8 | 2.61 | 0.01 | 0.69 |
| 7 | 38.0 | 12.7 | 33.2 | 13.7 | 2.4 | 2.69 | 0.96 | 0.70 |
| 8 | 42.5 | 17.1 | 24.9 | 8.3 | 7.3 | 2.70 | 0.10 | 0.71 |

### Exploratory factor analysis

After randomly assigning individuals to two groups (n = 130 for EFA and n = 120 for CFA), parallel analysis was used to determine the appropriate number of factors to retain. The initial analysis of the group 1 sample identified two factors with eigenvalues greater than one. The two-factor solution accounted for 79.09% of the total variance. The KMO measure was 0.71, and Bartlett’s test of sphericity was significant (χ^2^ = 1667.21; p < 0.001), indicating that the data was suitable for factor analysis (**Table 3**).

**Table 3.** Factor loading values for each item of the ISSS-8 and scale-level statistics (n= 130).

| ISSS-8 item | Factor 1 | Factor 2 |
| --- | --- | --- |
| Amongst the people you feel close to, is there someone who... | **Positive Support** | **Detrimental Interactions** |
| 1. Is there for you when you need him/her | 0.90 |  |
| 2. Gives you comfort | 0.77 |  |
| 3. Talks about important decisions with you | 0.90 |  |
| 4. Spends part of his/her time working some things out for you | 0.91 |  |
| 5. Worries too much about your illness |  | 0.91 |
| 6. Gives you information or makes suggestions that you find unhelpful or upsetting |  | 0.84 |
| 7. Makes you feel you cannot care for yourself |  | 0.95 |
| 8. Tries to change the way you're coping with your illness in a way you don't like |  | 0.88 |
| **Scale-level statistics** |  |  |
| KMO | 0.71 |  |
| Cronbach's alpha | 0.89 | 0.91 |
| Eigenvalue | 3.25 | 3.11 |
| % of variance | 52.34 | 26.75 |
| Cumulative % | 52.34 | 79.09 |

### Confirmatory factor analysis

To assess construct validity, a robust maximum likelihood CFA was conducted to fit the ISSS-8 eight-item measurement model. All eight ISSS-8 items were assigned to two factors consistent with the original structure of the ISSS-8: Positive Support (4 items) and Detrimental Interactions (4 items). A parallel examination of the factor components of the ISSS-8 was performed. The model demonstrated a reasonable fit to the data, as indicated by the five preset fit criteria (χ^2^/df = 1.585; p = 0.550, CFI = 0.99, AGFI = 0.97, TLI = 0.99, and RMSEA = 0.048 (95% CI = 0.023–0.051). All items in the model were significantly loaded onto their respective factors (all p-values < 0.05), except for the factor-constraint item, for which no significance test could be conducted. In addition, all factor loadings were greater than 0.50, indicating that individual ISSS-8 items were suitable for measuring the underlying construct of social support, albeit to varying degrees (Fig 1).

**Fig 1.**
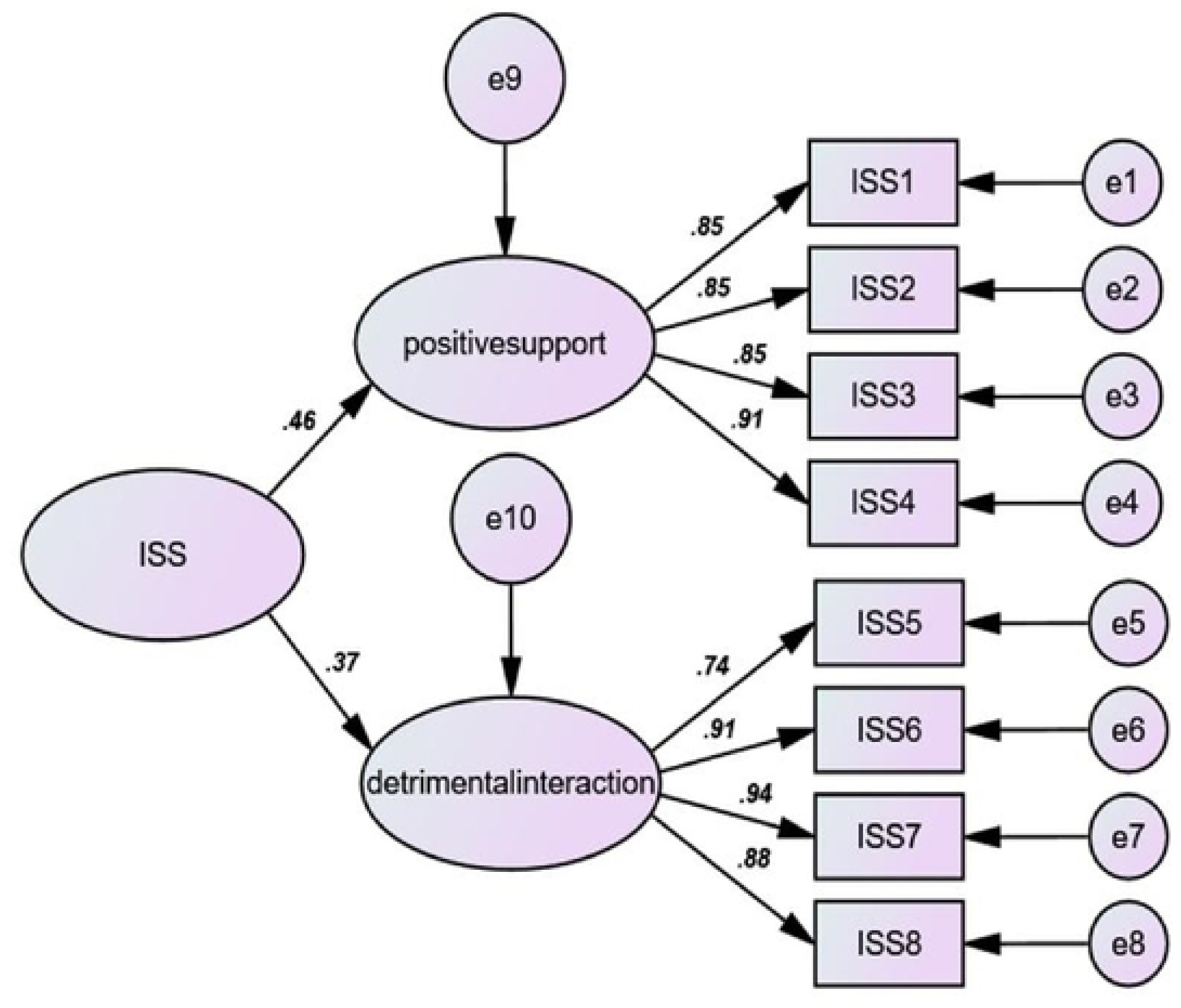
Structural model of the ISSS-8 showing standardized path coefficients.

### Reliability analysis

The CR for the two domains of the ISSS-8 ranged from 0.93 to 0.94, demonstrating acceptable reliability. The ISSS-8 also exhibited adequate internal consistency reliability, with Cronbach’s alpha values of 0.80 for the overall scale, 0.89 for Positive Support, and 0.91 for Detrimental Interactions. Strong internal consistency was supported by item–rest correlations for all items, ranging from 0.33 to 0.65, exceeding the recommended threshold of 0.30 (Table 4).

**Table 4.** Reliability analysis of the ISSS-8.

| Subscale | Number of items | CR | Cronbach's alpha | Item–rest correlation | AVE | ICC |
| --- | --- | --- | --- | --- | --- | --- |
| Positive Support | 4 | 0.93 | 0.89 | 0.33–0.45 | 0.76 | 0.924 |
| Detrimental Interactions | 4 | 0.94 | 0.91 | 0.56–0.65 | 0.80 | 0.933 |
| Overall |  |  | 0.80 |  |  |  |

### Convergent and discriminant validity

Table 4 presents the reliability analysis of the ISSS-8. AVE calculations revealed that all the ISSS-8 subscales exhibited convergent validity (Positive Support AVE = 0.76; Detrimental Interactions AVE = 0.80). The discriminant validity reflects the distinctiveness of the two subscales. The square root of AVE for Positive Support was 0.87, and for Detrimental Interactions it was 0.89, both exceeding the correlation coefficients between the subscales (r = 0.48, 0.37) (Figure 1). These findings confirm the presence of discriminant validity.

### Test–retest reliability

Test–retest reliability refers to the consistency of a measure when administered at different points in time. It is a critical aspect of evaluating the stability and dependability of assessment tools. During a two-week retest assessment, intraclass correlation coefficients (ICCs) were calculated for 30 participants, demonstrating adequate stability. The mean score on the social support scale decreased slightly from 16.16 (SD = 5.48) on Day 1 to 15.80 (SD = 5.13) on Day 14. The ICC value was 0.924 for Positive Support (95% CI = 0.918–0.952, p < 0.001), and 0.933 for Detrimental Interactions (95% CI = 0.913–0.962, p < 0.001).

### Concurrent validity

Concurrent validity of the ISSS-8 and its two subscales was assessed via correlation with the SICPA. As hypothesized, the total ISSS-8 scale and Detrimental Interactions subscale showed a statistically significant positive but small negative association with the total SICPA score, with correlations of -0.237 and 0.244, respectively. In addition, the Positive Support subscale showed a statistically significant association with the total ISSS-8 score (r = 0.634; p < 0.001), indicating a medium effect size. The correlation between Positive Support and Detrimental Interactions was low, supporting the notion that positive and negative interactions are independent of one another. (Table 5).

**Table 5.** Correlation between the ISSS-8 and concurrent measures.

|  | 1 | 2 | 3 | 4 |
| --- | --- | --- | --- | --- |
| 1. Positive Support | 1 |  |  |  |
| 2. Detrimental Interactions | 0.144 | 1 |  |  |
| 3. Total ISSS-8 | 0.672** | 0.836** | 1 | 0.244* |
| 4. Total SICPA | 0.113 | -0.237* | 0.244* | 1 |

\* All correlations are significant at a $p < 0.001$ level (two-tailed).

## Discussion

To the best of our knowledge, this is the first study to translate and validate the psychometric properties of the ISSS-8 for Thai patients with hematological malignancies in Northeastern Thailand. The ISSS-8 was effectively translated from English to Thai, employing a structured forward-and-back translation approach [24], with psychometric testing conducted following its translation and cross-cultural adaptation. The findings indicate that the ISSS-8 possesses acceptable psychometric properties and demonstrates validity for application in these patients, with participants reporting high levels of understanding and satisfaction. The examination of structural validity through EFA and CFA confirmed the presence of two factors within the ISSS-8 for this population: Positive Support (four items) and Detrimental Interactions (four items). These two factors are based on the factor structure of the German version of the ISSS-8, as described by Ramm and Hasenbring [16]. While the Positive Support Subscale measures advantageous and encouraging interactions, such as feeling understood, receiving emotional comfort, and getting practical help, the Detrimental Interactions Subscale measures problematic or unhelpful interactions, such as overprotectiveness, feeling criticized, or receiving excessive pessimism/optimism. Hence, construct validity of the ISSS-8 was supported by CFA, with all fit indices indicating acceptable model fit, supporting the two-dimensional structure of the ISSS-8 as a satisfactory fit for the data. Although a substantial chi-square was observed, all items exhibited significant standardized loadings, thereby affirming their validity in representing the intended constructs [32]. The findings collectively endorse the ISSS-8 as an effective instrument for evaluating social support among patients with hematological malignancies. These results are consistent with previous studies validating this tool in patients with cancer or other chronic illnesses, predominantly in Western contexts [15–17].

The results of this study showed ceiling effects on all items of the Positive Support Subscale and floor effects on all items of the Detrimental Interactions Subscale. The ceiling effect in the Positive Support Subscale indicates that the distribution of responses is heavily skewed towards the maximum possible score [28]. The ISSS-8 was designed to assess social support, a factor prevalent in the majority of the population, including those with hematological malignancies. However, the sensitivity of this subscale is reduced because it becomes difficult for the Positive Support Subscale to detect improvements in social support over time, such as after a psychological intervention, if the baseline score is already near the maximum. Likewise, floor effects existed on the Detrimental Interactions Subscale. This finding may be due to the fact that most participants do not experience high levels of explicitly critical or detrimental interactions related to their illness. Therefore, a large percentage of respondents score at or near the minimum possible score [28]. In addition, the items of each scale exhibited strong discrimination indices. All item-total correlations exhibited a strong positive association with the total score, indicating the scale’s homogeneity. These indices suggest that the Thai-version items of the ISSS-8 could effectively differentiate between high and low scores on this scale. This result is consistent with a study exploring illness-specific positive social support and detrimental interactions in melanoma survivors [17].

The reliability of the ISSS-8 was verified using internal consistency and a test–retest technique. The CR of the two domains of the ISSS-8 varied from 0.93 to 0.94, signifying excellent reliability. The Cronbach’s alpha values in our study indicated that the ISSS-8 total scores and its subscales exhibited strong internal consistency [34]. The high internal consistency indicates that respondents who select high scores for one item are likely to select high scores for other items as well, and conversely. The findings align with results from various validation studies conducted across diverse populations, including cancer and other chronically ill patients [15–17, 38]. However, these findings are inconsistent with results from a prior study on samples of cancer patient–relative dyads, which reported that internal consistency was partly lower but still acceptable [39]. Furthermore, the item analysis results indicated that the items of each scale exhibited strong discrimination indices. All item–total correlations exhibited a strong positive association with the total score, indicating the scale’s homogeneity. The indices suggest the potential to differentiate between high and low scores on this scale utilizing the Thai-version items of the ISSS-8. The tool’s strong reliability indicates it is well-designed and able to respond efficiently to inquiries, as demonstrated by participant feedback. This evidence confirms that the ISSS-8 is well-regarded among patients with hematological malignancies in Thailand. Related research indicates that this assessment instrument provides a high item discrimination index [17]. Moreover, test–retest reliability was assessed over a two-week period with a sample of 30 participants utilizing ICC. The findings exhibited adequate stability, evidenced by an ICC of 0.924 for the Positive Support domain and 0.933 for Detrimental Interactions, signifying excellent dependability in accordance with established standards [34]. The convergent validity of the ISSS-8 was assessed by calculating the AVE for both subscales. The findings indicated that the AVE for all subscales was above 0.50, exceeding the recommended threshold for convergent validity. This approach aligns with recent psychometric validations of perceived social support measures among people with dementia [40]. Additionally, the ISSS-8 exhibited robust discriminant validity per the Fornell-Larcker standard [37], as the square root of the AVE surpassed the correlation between the ISSS-8 and the SICPA. This means that the ISSS-8 detects differences that the social participation measure does not. These findings, along with the high item–rest correlations, provide strong evidence of validity based on the internal structure and interrelations of the variables.

For concurrent validity, as hypothesized, the total ISSS-8 scale and Detrimental Interactions subscale showed a small, positive, statistically significant association with the total score of SICPA measures. The results likely reflect that the SICPA assesses self-efficacy (the "I can accomplish this" aspect). A patient may encounter "detrimental interactions" (such as a spouse exhibiting excessive emotional distress or criticism) yet interpret it as "They are merely frightened" rather than "I am inadequate." This result is consistent with previous research using the ISSS-8, which confirms that while positive support is high, "detrimental interactions" are significantly linked to higher distress (SICPA-related constructs) [17]. In addition, self-efficacy denotes a patient’s belief in their capacity to manage cancer, cope with treatment side effects, and address the emotional challenges associated with the disease. Research consistently demonstrates a negative correlation between detrimental interactions and self-efficacy [18].

### Strengths and limitations

In this study, we utilized an adequate sample size to analyze the psychometrics of the Thai version of the ISSS-8, alongside a small number of participants to evaluate the test–retest reliability. However, specific limitations that may have impacted the findings must be recognized. Firstly, the sample was sufficient for an initial validation study; nevertheless, it was restricted to three tertiary hospitals in Northeastern Thailand, and the use of convenience sampling in clinical settings may limit the generalizability of the research findings to other populations, particularly in areas with notable cultural or linguistic diversity. Future research may employ alternative approaches, such as stratified sampling across national tertiary hospitals, to obtain more diverse and inclusive samples for assessing the psychometric validation of the Thai version of the ISSS-8. Second, our study used a self-report questionnaire, which has inherent limitations. Response bias may have affected data accuracy. To mitigate this issue, all participants were assured of the anonymity of their responses. They were instructed to respond honestly and not share their answers with others. Finally, the study primarily focused on participants without mental illnesses. Future research should expand the scope to include individuals with pre-existing conditions, such as anxiety and depression.

## Conclusion

This research translated and validated the psychometric properties of the Thai version of the ISSS-8, establishing it as a reliable instrument for assessing social support in patients with hematological malignancies. Substantial evidence was obtained to confirm its reliability, convergent validity, and construct validity within this population, as verified through CFA and supporting statistical evaluations. The instrument effectively captures all dimensions of social support within this population, closely resembling the original ISSS-8 while incorporating culturally relevant adaptations. Its demonstrated validity and reliability highlight its utility for healthcare professionals in assessing social support levels in patients with hematological malignancies in both research and clinical contexts. These findings emphasize the importance of integrating social support assessments into cancer management and family-centered care, thereby enhancing holistic and responsive healthcare delivery in Thailand.

## Data Availability

All data produced in the present study are available upon reasonable request to the authors.

## Acknowledgements

The authors would like to thank Dr Gesa C. Ramm for granting permission to use the ISSS-8. The authors sincerely thank all the patients with hematological malignancies who participated in this study. We greatly appreciate their willingness to provide personal information for the purposes of this research.

## Supporting information

ISSS1 data.

(XLSX)

## Author contributions

**Conceptualization:** Ueamporn Summart, Monthida Sangruangake

**Data curation:** Monthida Sangruangake, Chaliya Wamaloon, Acharaporn Moungmultri

**Formal analysis:** Ueamporn Summart, Muhamad Zulfatul A’la

**Methodology:** Ueamporn Summart, Monthida Sangruangake

**Supervision:** Ueamporn Summart

**Visualization:** Ueamporn Summart, Monthida Sangruangake

**Writing – original draft:** Ueamporn Summart, Monthida Sangruangake, Chaliya Wamaloon, Acharaporn Moungmultri, Muhamad Zulfatul A’la

**Writing –review&editing:** Ueamporn Summart, Monthida Sangruangake, Chaliya Wamaloon, Acharaporn Moungmultri, Muhamad Zulfatul A’la

**Competing interests:** The authors declare they have no competing interests.

## Notes

### Competing Interest Statement

The authors have declared no competing interest.

### Funding Statement

This study did not receive any funding.

### Author Declarations

Ethics Committee of Udon Thani Rajabhat University gave ethical approval for this work.

## References

1. Zhang N, Wu J, Wang Q, Liang Y, Li X, Chen G, et al. Global burden of hematologic malignancies and evolution patterns over the past 30 years. Blood Cancer Journal. 2023;13(1):82. 10.1038/s41408-023-00853-3. PMID: 37193689

2 Mitchell E, Pham MH, Clay A, Sanghvi R, Williams N, Pietsch S, et al. The long-term effects of chemotherapy on normal blood cells. Nature genetics. 2025;57(7):1684–94. 10.1038/s41588-025-02234-x. PMID: 40596443

3. Bellali T, Manomenidis G, Meramveliotaki E, Minasidou E, Galanis P. The impact of anxiety and depression in the quality of life and psychological well-being of Greek hematological cancer patients on chemotherapy. Psychology, health & medicine. 2020;25(2):201–13. 10.1080/13548506.2019.1695864. PMID: 31777270

4. Billingy NE, Tromp VN, Veldhuijzen E, Belderbos J, Aaronson NK, Feldman E, et al. Symptom monitoring with Patient-Reported Outcomes using a web application among patients with Lung cancer in the Netherlands (SYMPRO-Lung): study protocol for a stepped-wedge randomised controlled trial. Bmj Open. 2021;11(9):e052494. doi:10.1136/bmjopen-2021-052494. PMID: 34518276

5. Daré LO, Bruand P-E, Gérard D, Marin B, Lameyre V, Boumédiène F, et al. Co-morbidities of mental disorders and chronic physical diseases in developing and emerging countries: a meta-analysis. BMC public health. 2019;19(1):304. doi: 10.1186/s12889-019-6623-6. PMID: 30866883

6. Kim ES, Tkatch R, Martin D, MacLeod S, Sandy L, Yeh C. Resilient aging: Psychological well-being and social well-being as targets for the promotion of healthy aging. Gerontology and geriatric medicine. 2021;7:23337214211002951. doi: 10.1177/23337214211002951. PMID: 33816707

7. Sanna K, Cierpiałkowska L, Kleka P, Stelmach-Mardas M, Iżycki D. Development of the cancer-patient social support questionnaire: reliability and validity. 2019.

8. Lin Y, He M, Zhou W, Zhang M, Wang Q, Chen Y, et al. The relationship between physical exercise and psychological capital in college students: the mediating role of perceived social support and self-control. BMC Public Health. 2025;25(1):581. doi: 10.1186/s12889-025-21856-8. PMID: 39939931

9. Köhler N, Mehnert A, Götze H. Psychological distress, chronic conditions and quality of life in elderly hematologic cancer patients: study protocol of a prospective study. BMC cancer. 2017;17(1):700. doi: 10.1186/s12885-017-3662-1. PMID: 29070033

10. Zhu J, Sjölander A, Fall K, Valdimarsdottir U, Fang F. Mental disorders around cancer diagnosis and increased hospital admission rate-a nationwide cohort study of Swedish cancer patients. BMC cancer. 2018;18(1):322. doi: 10.1186/s12885-018-4270-4. PMID: 29580232.

11. Corovic S, Vucic V, Mihaljevic O, Djordjevic J, Colovic S, Radovanovic S, et al. Social support score in patients with malignant diseases—with sociodemographic and medical characteristics. Frontiers in Psychology. 2023;14:1160020. doi: 10.3389/fpsyg.2023.1160020. eCollection 2023. PMID: 37325739.

12. Wongpakaran N, Wongpakaran T. A revised Thai multi-dimensional scale of perceived social support. The Spanish journal of psychology. 2012;15(3):1503–9. doi:10.5209/rev_sjop.2012.v15.n3.39434. PMID: 23156952.

13. Wongpakaran T, Wongpakaran N, Sirirak T, Arunpongpaisal S, Zimet G. Confirmatory factor analysis of the revised version of the Thai multidimensional scale of perceived social support among the elderly with depression. Aging & mental health. 2018;22(9):1149–54.

14. Jaiboon S, Vittayatigonnasak A. Factors Influencing the Quality of Life in Thai Cancer Patients Receiving Chemotherapy during COVID-19 Pandemic. 2023. doi:10.1080/13607863.2017.1339778. PMID: 28621147

15. Ullrich A, Mehnert A. Psychometrische evaluation and validierung einer 8-Item Kurzversion der Skalen zur Sozialen Unterstützung bei Krankheit (SSUK) bei Krebspatienten. Klinische Diagnostik und Evaluation. 2010;3(359):81.

16. Ramm GC, Hasenbring M. Die deutsche Adaptation der Illness-specific Social Support Scale und ihre teststatistische Überprüfung beim Einsatz an Patienten vor und nach Knochenmarktransplantation. Zeitschrift für Medizinische Psychologie. 2003;12(1):29–38. 10.3233/ZMP-2003-12_1_06

17. Fischbeck S, Weyer-Elberich V, Zeissig SR, Imruck BH, Blettner M, Binder H, et al. Determinants of illness-specific social support and its relation to distress in long-term melanoma survivors. BMC public health. 2018;18(1):511.

18. Geue K, Götze H, Friedrich M, Leuteritz K, Mehnert-Theuerkauf A, Sender A, et al. Perceived social support and associations with health-related quality of life in young versus older adult patients with haematological malignancies. Health and Quality of Life Outcomes. 2019;17(1):145. doi: 10.1186/s12955-019-1202-1. PMID: 31438983

19. Timko Olson ER, Olson A, Driscoll M, Bliss DZ. Psychosocial Factors Affecting Wellbeing and Sources of Support of Young Adult Cancer Survivors: A Scoping Review. Nursing Reports. 2024;14(4):4006–21.doi:10.3390/nursrep14040293. PMID: 39728654

20. Kardosod A, Needham J, Udomkhwamsuk W, Wongcharoen W, Petsky H. Experiences, Coping and Caregiver Well-being for Caregiving Thai Family Members with Metastatic Spinal Cancer for Palliative Home Care: A Descriptive Qualitative Study. European Journal of Oncology Nursing. 2025:102919. doi: 10.1016/j.ejon.2025.102919. PMID: 40592233

21. Posai V, Srisintorn W, Thongsuksai P, Jitpanya C. Psychometric Evaluation of The Thai Version of Emotional Labor Scale (T-ELS) in Thai Nurses. Journal of Health Research. 2025;39(3):10. https://digital.car.chula.ac.th/jhr

22. Bontempo AC, Bontempo JM, Duberstein PR. Ignored, dismissed, and minimized: Understanding the harmful consequences of invalidation in health care—A systematic meta-synthesis of qualitative research. Psychological Bulletin. 2025;151(4):399. doi: 10.1037/bul0000473. PMID: 40310228

23. Cain CL, Surbone A, Elk R, Kagawa-Singer M. Culture and palliative care: preferences, communication, meaning, and mutual decision making. Journal of pain and symptom management. 2018;55(5):1408–19. doi: 10.1016/j.jpainsymman.2018.01.007. PMID: 29366913

24. Jones PS, Lee JW, Phillips LR, Zhang XE, Jaceldo KB. An adaptation of Brislin’s translation model for cross-cultural research. Nursing research. 2001;50(5):300–4. doi: 10.1097/00006199-200109000-00008. PMID: 11570715

25. Revenson TA, Schiaffino KM, Majerovitz SD, Gibofsky A. Social support as a double-edged sword: The relation of positive and problematic support to depression among rheumatoid arthritis patients. Social science & medicine. 1991;33(7):807–13. doi: 10.1016/0277-9536(91)90385-p. PMID: 1948172

26. Telch CF, Telch MJ. Group coping skills instruction and supportive group therapy for cancer patients: a comparison of strategies. Journal of consulting and clinical psychology. 1986;54(6):802. doi: 10.1037//0022-006x.54.6.802. PMID: 3794024

27. Kline RB. Principles and practice of structural equation modeling: Guilford publications; 2023.

28. Terwee CB, Bot SD, de Boer MR, Van der Windt DA, Knol DL, Dekker J, et al. Quality criteria were proposed for measurement properties of health status questionnaires. Journal of clinical epidemiology. 2007;60(1):34–42. doi: 10.1016/j.jclinepi.2006.03.012. PMID: 17161752

29. Schmeiser CB, Welch CJ. Test development. Educational measurement. 2006;4:307–53.

30. Kaiser HF. An index of factorial simplicity. psychometrika. 1974;39(1):31–6.

31. Hu Lt, Bentler PM. Cutoff criteria for fit indexes in covariance structure analysis: Conventional criteria versus new alternatives. Structural equation modeling: a multidisciplinary journal. 1999;6(1):1–55. 10.1080/10705519909540118

32. Satorra A, Bentler PM. A scaled difference chi-square test statistic for moment structure analysis. Psychometrika. 2001;66(4):507–14. 10.1007/BF02296192

33. Kline P. Handbook of psychological testing: Routledge; 2013.

34. Streiner DL, Norman GR, Cairney J. Health measurement scales: a practical guide to their development and use: Oxford university press; 2024. doi: 10.1111/1753-6405.12484. PMID: 27242256

35. Fayers PM, Machin D. Quality of life: the assessment, analysis and reporting of patient-reported outcomes: John Wiley & Sons; 2015.

36. Cohen J. Statistical power analysis for the behavioral sciences: routledge; 2013. 10.4324/9780203771587

37. Fornell C, Larcker DF. Evaluating structural equation models with unobservable variables and measurement error. Journal of marketing research. 1981;18(1):39–50. 10.2307/3151312.

38. Müller D, Mehnert A, Koch U. Skalen zur Sozialen Unterstützung bei Krankheit (SSUK)–Testtheoretische Überprüfung und Validierung an einer repräsentativen Stichprobe von Brustkrebspatientinnen. Zeitschrift für medizinische Psychologie. 2004;13(4):155–64.

39. Hermann M, Goerling U, Hearing C, Mehnert-Theuerkauf A, Hornemann B, Hövel P, et al. Social Support, Depression and Anxiety in Cancer Patient-Relative Dyads in Early Survivorship: An Actor-Partner Interdependence Modeling Approach. Psycho-Oncology. 2024;33(12):e70038. doi: 10.1002/pon.70038.

40. Vidyanti AN, Nafiati R, Putri GFS, Prodjohardjono A, Effendy C. Measurement of perceived social support among people with dementia: a validation of the Indonesian version of the personal resource questionnaire-2000 (PRQ2000-INA). BMC geriatrics. 2025;25(1):1–11. doi: 10.1186/s12877-025-06473-9. PMID: 41220032.

